# A collaborative maternity and newborn dashboard (CoMaND) for the COVID-19 pandemic: a protocol for timely, adaptive monitoring of perinatal outcomes in Melbourne, Australia

**DOI:** 10.1101/2021.07.22.21261008

**Authors:** Lisa Hui, Melvin Barrientos Marzan, Stephanie Potenza, Daniel L. Rolnik, Joanne M. Said, Kirsten R Palmer, Clare L. Whitehead, Penelope M. Sheehan, Jolyon Ford, Natasha Pritchard, Ben W. Mol, Susan P. Walker

## Abstract

**Background:** The COVID-19 pandemic has resulted in a range of unprecedented disruptions to the delivery of maternity care globally and has been associated with regional changes in perinatal outcomes such as stillbirth and preterm birth. Metropolitan Melbourne endured one of the longest and most stringent lockdowns in 2020. This paper presents the protocol for a collaborative maternity dashboard project to monitor perinatal outcomes in Melbourne, Australia, during the COVID-19 pandemic.

**Methods:** De-identified maternal and newborn outcomes will be collected monthly from all public maternity services in Melbourne, allowing rapid analysis of a multitude of perinatal indicators. Weekly outcomes will be displayed as run charts according to established methods for detecting non-random ‘signals’ in health care. A pre-pandemic median for all indicators will be calculated for the period of January 2018 to March 2020. A significant shift is defined as ≤ six consecutive weeks, all above or below the pre-pandemic median. Additional statistical analyses such as regression, time-series, and survival analyses will be performed for an in-depth examination of maternal and perinatal outcomes of interests.

**Ethics and Dissemination:** This study has been registered as an observational study with the Australian and New Zealand Clinical Trials Registry (ACTRN12620000878976).

**Strengths and weaknesses of this study:**

⍰ This project is the first clinician-led, multi-centre perinatal data collection system for metropolitan Melbourne.
⍰ It complements the state government data collection, with the significant benefits of more timely and flexible reporting of outcomes, and granular detail on emerging areas of concern.
⍰ The study relies on primary source coding of exposure and outcomes from each hospital that have not been internally validated during the study period.
⍰ Data from private maternity hospitals, containing 25% of Melbourne births, are not available.
⍰ This resource will support data-informed hospital pandemic responses through to the end of 2022.

## BACKGROUND

The COVID-19 pandemic has resulted in a range of unprecedented disruptions to the delivery of maternity care globally. International studies have reported significant changes in perinatal outcomes such as stillbirths and preterm birth during the COVID-19 pandemic, raising concerns about the unintended consequences of pandemic restrictions on mothers and newborns [1-4]. Women in metropolitan Melbourne endured one of the longest and most stringent pandemic lockdowns in the world (stringency index 92.6/100 for approximately 6 weeks), while continuing to give birth to ≤ 4000 babies per month ([5]) (See figure 1 and supplemental data 1).

**Figure 1.**
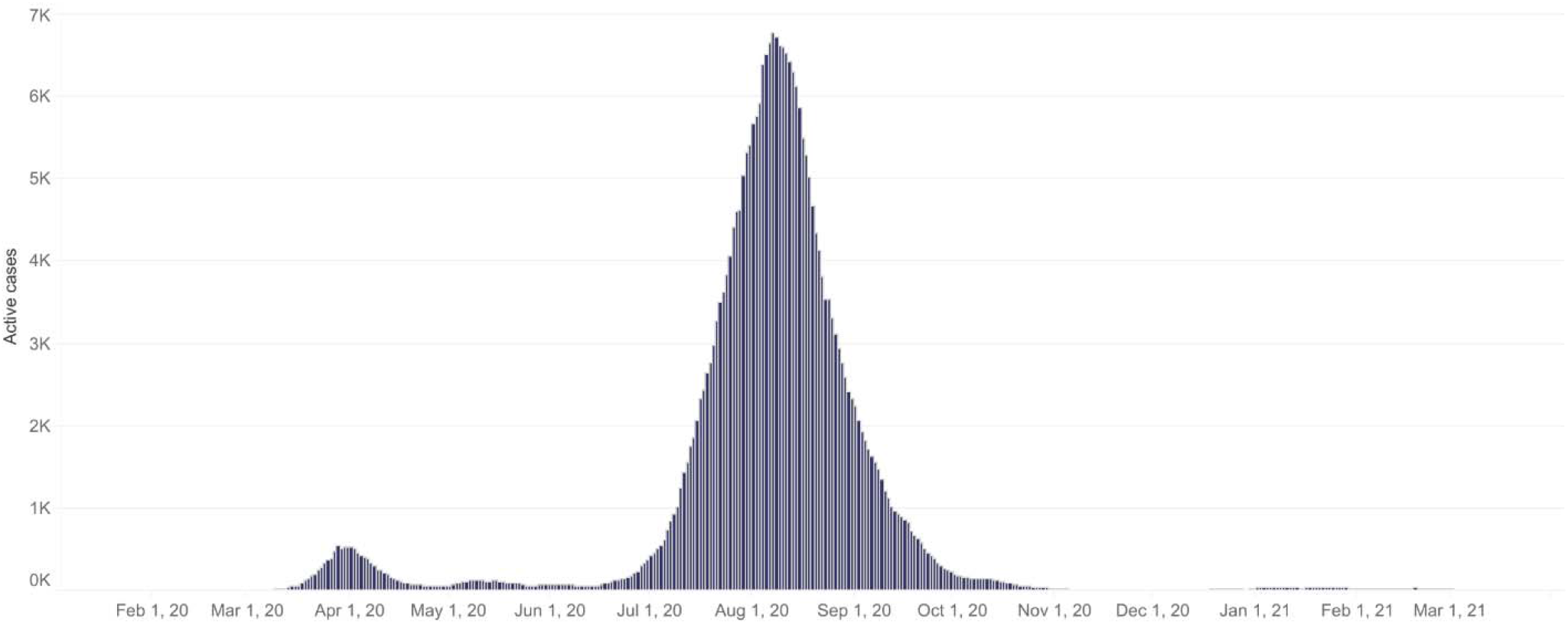
Number of active coronavirus cases by month in Victoria, Australia. Image downloaded from the Victorian Department of Health and Human Services at https://www.dhhs.vic.gov.au/victorian-coronavirus-covid-19-data on 15th March 2021

In April 2020, the Royal Australian College of Obstetricians and Gynaecologists (RANZCOG) released a communique outlining suggested changes to routine pregnancy care to mitigate the anticipated strain on health services and infection risk to patients and health care workers [6].

These measures included:

⍰ Reducing, postponing and/or increasing the interval between antenatal visits
⍰ Limiting the time of all antenatal visits to less than 15 minutes
⍰ Using telehealth consultations as a replacement, or in addition to, routine visits
⍰ Cancelling face to face antenatal classes
⍰ Limiting visitors (partner only) while in hospital
⍰ Considering early discharge from hospital
⍰ Modifications to screening for gestational diabetes

At the time of implementation, the impact of these widespread changes to antenatal care was unknown. Victorian maternity services collect a comprehensive set of data to fulfil legislative and regulatory requirements. These data are submitted to Safer Care Victoria (SCV) for the biannual Mother’s and Babies’ Report and the Perinatal Services Performance Indicator (PSPI) report [7]. However, there is typically a minimum 18-month lag time between data submission and report publication.

The primary aim of this study is to leverage routinely collected data from all public maternity units in Melbourne to create a timely, flexible monitoring system to monitor perinatal outcomes during the COVID-19 pandemic. Weekly trends in key maternal and newborn outcomes before, during and after the COVID-19 pandemic will be analysed through monthly data uploads from participating health services. The initial study period will include two years preceding the WHO declaration of the “Public Health Emergency of International Concern” in January 2020 [8] for baseline pre-pandemic rates of key performance indicators. As there is uncertainty around the duration of the pandemic, the study will extend to at least 31^st^ December 2022 to account for the length of gestation, lag effects, COVID-19 vaccination roll-out and ongoing alterations to maternity care through 2020 and 2021. The secondary aim of this study is to investigate the effect of lockdowns on outcomes of interest, including preterm birth, stillbirth, and birth weight.

## METHODS

### Study Design

This multi-centre retrospective cohort study will gather routinely collected data on births in public maternity hospitals in Victoria from 1^st^ January 2018, to 31^st^ December 2022. The first half of the study period (from 1^st^ January 2018 to commencement of study following ethics and local governance approvals in mid-2020) will be retrospectively collected to provide baseline pre-pandemic medians rates of all outcomes. Data will then be prospectively collected each calendar month from each of the participating sites, and this will be updated to provide regular reports on maternity and perinatal services.

### Participating health services

The participating health services and contributing hospitals are:

⍰ Mercy Health (Mercy Hospital for Women, Werribee Mercy Hospital)
⍰ The Royal Women’s Hospital, The Women’s at Sandringham
⍰ Monash Health (Monash Medical Centre, Casey Hospital, Dandenong Hospital)
⍰ Northern Health (The Northern Hospital)
⍰ Western Health (Joan Kirner Women’s and Children’s Hospital)
⍰ Eastern Health (Box Hill Hospital, The Angliss Hospital)
⍰ Peninsula Health (Frankston Hospital)

These seven health services (12 hospitals) include four level 6 maternity centres, three-level 5 centres and five-level 4 health services. Level 4 centres provide local care for low-risk mothers and babies including births from 34 weeks’ gestation. Levels 5 centres provide care for women with normal to moderate risk pregnancies, including births from 31 weeks’ gestation. Level 6 centres provide comprehensive maternity care for women of any risk level, including maternal-fetal medicine services and level 6 newborn care for babies < 31 weeks’ gestation. The detailed definitions of each service capability level are described elsewhere [9].

### Study population

All births ≤ 20 weeks with babies with a birth weight ≤150 grams recorded at one of the participating maternity hospitals between 1^st^ January 2018 and 31^st^ December 2022 will be included. Birth < 20 weeks gestation are excluded as these data are not captured in the current maternity data collection systems. An estimated 44,000 women give birth in the participating hospitals each year [10]; the number of births captured during the full study period (2018-2022) will be approximately 220,000. Data from private hospitals will not be collected. Our participating sites will therefore capture approximately 74% of all hospital births in metropolitan Melbourne, including the highest risk pregnancies requiring referral to level 6 centres (Table 1).

**Table 1.**
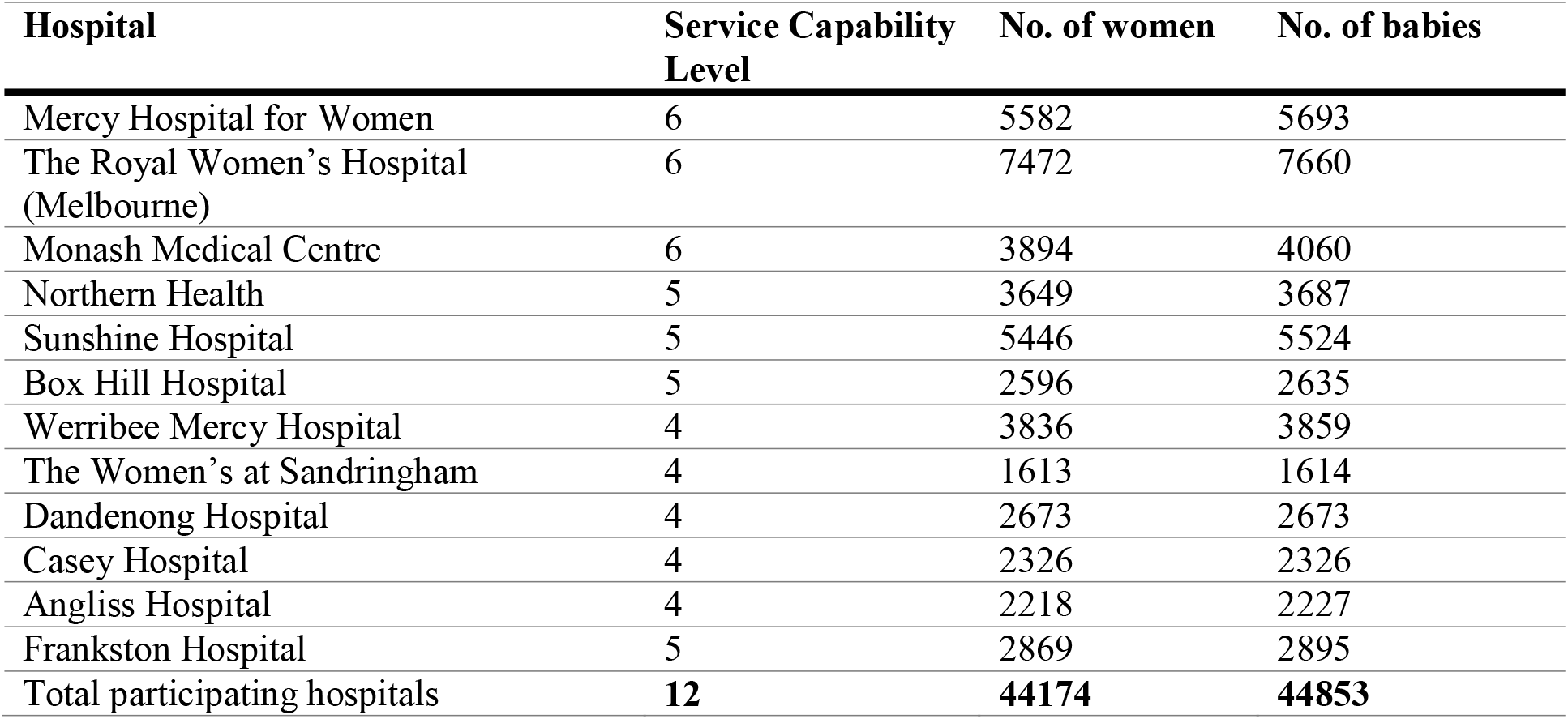
Melbourne births by hospital in 2018. (Hunt, Davey et al. 2019)

| **Hospital** | **Service Capability Level** | **No. of women** | **No. of babies** |
| --- | --- | --- | --- |
| Mercy Hospital for Women | 6 | 5582 | 5693 |
| The Royal Women's Hospital (Melbourne) | 6 | 7472 | 7660 |
| Monash Medical Centre | 6 | 3894 | 4060 |
| Northern Health | 5 | 3649 | 3687 |
| Sunshine Hospital | 5 | 5446 | 5524 |
| Box Hill Hospital | 5 | 2596 | 2635 |
| Werribee Mercy Hospital | 4 | 3836 | 3859 |
| The Women's at Sandringham | 4 | 1613 | 1614 |
| Dandenong Hospital | 4 | 2673 | 2673 |
| Casey Hospital | 4 | 2326 | 2326 |
| Angliss Hospital | 4 | 2218 | 2227 |
| Frankston Hospital | 5 | 2869 | 2895 |
| Total participating hospitals | **12** | **44174** | **44853** |

The age profile of mothers in Victoria is similar to the Australian average, with 38% of women giving birth being aged 30-34 years old [11].

### Data Sources

There are two sources of data.

### 1. Maternity hospital data collection systems

Perinatal outcomes will be collected from the electronic Birthing Outcomes System (BOS, version 6·04, Management Consultants and Technology Services, Caulfield, Victoria, Australia. https://www.mcats.com.au/) (or equivalent) at each participating hospital. Each BOS site will house identical embedded report queries that will be run monthly with minimal resource requirements (less than one hour of work per month to generate output). No individual identifying data (e.g., date of birth, maternal date of birth, medical record number) will be collected for this study.

A comprehensive set of variables are being collected to enable granular reporting on outcomes, including, but not limited to, those listed below. The complete list of variables is listed in Appendix 2. As this study has a flexible, adaptive design, additional outcomes can be generated from the list of collected variables as required. The key minimum indicators are listed below:

Health System Outcomes

⍰ Total births per week
⍰ Total pregnancies per week

Maternal Outcomes

⍰ First antenatal visit
⍰ Gestational diabetes
⍰ Multiple pregnancies
⍰ Maternal body mass index
⍰ Labour onset
⍰ Mode of birth
⍰ Planned setting of birth
⍰ Maternal influenza vaccination rates
⍰ Smoking cessation
⍰ Perineal tears for vaginal births
⍰ General Anaesthesia for caesarean births
⍰ Length of stay
⍰ Post-partum haemorrhage

Fetal or Neonatal Outcomes

⍰ Stillbirth rates (unadjusted)
⍰ Preterm births
⍰ birth weight <= 3^rd^ centile
⍰ Rate of Apgar scores < 7 at 5 minutes of age
⍰ Breastfeeding rates
⍰ Macrosomia (birthweight ≤=4000 grams and ≤=90^th^ centile)
⍰ Admission to Neonatal Intensive Care Unit (NICU) or Special Care Nursery (SCN)

### 2. State-wide billing for relevant Medicare Benefits Schedule (MBS) item numbers

The quarterly number of antenatal and postnatal consultations performed by a doctor, nurse, midwife or an Aboriginal and Torres Strait Islander health practitioner will be estimated from MBS statistics available at the Australian Department of Health website (http://medicarestatistics.humanservices.gov.au/statistics/mbs_item.jsp). We will also collect MBS item numbers for COVID-19 Temporary MBS Telehealth services and obstetric ultrasound billing. See appendix 3 for details.

## DATA COLLECTION PROCEDURES

### Description of procedures

1. During the third week of each month, perinatal outcomes for the preceding calendar month will be extracted from the Birthing Outcomes System (or equivalent) by site investigators at each health service. The data will be deidentified and uploaded to a secure online REDCap database administered by the University of Melbourne.
2. MBS item numbers for Victoria by quarter will be downloaded from the Australian Department of Health website and collated by the central study team. http://medicarestatistics.humanservices.gov.au/statistics/mbs_item.jsp
3. Once complete, the report will be disseminated to all investigators by email.

The CoMaND project is designed to be adaptive to rapid changes in the healthcare environment. Therefore, the reported indicators may be adjusted during the course of the study in order to respond to potential areas of concern (e.g., emerging high-risk groups that require specific monitoring).

### Data collection and management

The coordinating principal investigator (CPI) will be responsible for storing essential study documents relevant to central data management and maintaining site-specific data submission records. A data collection log and instruction manual will be maintained to record all handling of submitted data from each site. The central study team will merge and analyse the centralised data for the report.

Study data will be collected and managed using REDCap electronic data capture tools [12] hosted at The University of Melbourne. Data is backed up nightly to a local backup server. REDCap maintains an audit trail of data create/update/delete events that is accessible to project users who are granted permission to view it. Access to REDCap will be provided via The University of Melbourne user account or (for external collaborators) via a REDCap user account created by the University of Melbourne system administrator. Working files for central data clean up, merging, analysis and storage will be held on password-protected computers and archived on University of Melbourne servers.

### Statistical methods

Statistical analyses will be performed on the data collected from the CoMaND reports in two steps. (1) Analyses for routine clinical quality reporting (2) Statistical analyses for research purposes.

#### (1) Analyses for clinical quality reporting

The primary aim of this data collection is to detect any significant deviations in performance indicators over time, relative to the baseline pre-pandemic medians. The pre-pandemic medians will be calculated from the period encompassing 1 January 2018 to 31 March 2020 (onset of stage 3 lockdown). Deidentified individual patient-level data on ≤ 100 fields will be analysed in STATA SE v16 [13]. Weekly outcomes will be displayed as run charts according to established methods for detecting non-random ‘signals’ in health care as described elsewhere [14]. Interpretations will be made according to probability-based rules for non-random patterns of data (alpha error of < 0.05). The signal will be categorised as “shift” if there are ≤ 6 consecutive weeks all above or below the pre-pandemic median. A “trend” is defined as ≤ 5 consecutive weeks continuously going all up or all down. Detecting a signal suggesting deviation from a stable baseline will prompt communication to participating health services to allow further scrutiny for possibly contributory factors and areas for improvement.

#### (2) Statistical analyses for research purposes

Further in-depth analyses will be performed (i.e., survival analysis for pre-term births, regression analyses for macrosomia, time-series analyses for all other outcomes etc.) to determine the effects of lockdowns on the outcomes of interests. We will compare the maternal, perinatal, and neonatal outcomes between the cohort of women who were pregnant between weeks 20-37 gestational age during the period of most stringent pandemic restrictions in metropolitan Melbourne from March 31 2020 to November 23 2020.

The exposure and control groups were defined as the following:

⍰ Pandemic exposed cohort
  - Pregnancies with conception during the period commencing week of 25/11/2019 to the week of 23/03/2020 inclusive.
⍰ Comparison group (non-pandemic exposed)
  - Pregnancies from same calendar periods 12 and 24 months prior.

The outcomes to be included are preterm births (iatrogenic and spontaneous), stillbirths, antenatal corticosteroid use, Apgars <5, admission to Neonatal ICU or Special care nursery, Intrauterine growth restrictions (IUGR), and fetal macrosomia. Week of conception will be calculated from week of birth and weeks of gestational age at birth. Week of conception is computed using this formula:

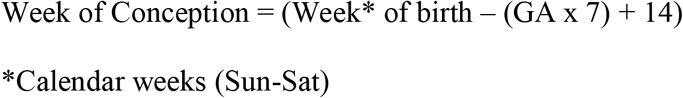

All pregnancies resulting to a singleton birth of at least 20 weeks gestation will be included whereas those with major anomalies and terminated pregnancies will be excluded.

Various covariates will also be used to adjust the multivariate analyses performed on each maternal and perinatal outcome. These covariates include: maternal age (<25, 25-29, 30-34, 35-39, 40+), region of birth (Australian Bureau of Statistics classifications), interpreter required (yes, no), parity (0,1, 2, ≤=3), maternal body mass index, socioeconomic Indexes for Areas (SEIFA, 2016): index of relative socioeconomic advantage and disadvantage (IRSAD) scores for the maternal postcode of residence, smoking status.

### Report dissemination

The reports will be provided to the following people listed below each month or as otherwise deemed practicable.

⍰ CPI and site PIs
⍰ Chair of the Melbourne Academic Centres for Health, Women’s and Newborn network
⍰ Senior maternity advisor to Safer Care Victoria / Chair of Consultative Council of Obstetric and Paediatric Morbidity and Mortality, who will disseminate to the members of the COVID-19 expert obstetric advisory group to Safer Care Victoria

Authorised representatives of the sponsoring institution as well as representatives from the human research ethics committees, research governance office and regulatory agencies may inspect all documents and records required to be maintained by the CPI in this study.

In addition to the regular report circulation as described above, results may also be written up for journal publication and/or presentation at scientific conferences. Wherever possible, research publications will be published under an open access policy. Data sharing with international investigators will be in accordance with relevant ethics approvals. The results will be distributed to the relevant clinical service leaders, who will translate the findings into clinical guidelines and protocols as appropriate.

### Data confidentiality

No individually identifying data will be collected or stored. Outcomes will be reported in aggregate forms, and results will not be disseminated in a format that allows the identification of an individual patient.

### Archiving - Data and document retention

Data will be stored indefinitely. The CPI will be the custodian of the data. Local site study documents will be stored for at least 15 years and will not be destroyed without the written consent of the CPI. The CPI will inform Principal Site Investigators when these documents no longer need to be retained. Secure destruction of research documents using software to permanently erase the data will be utilised at the end of the archive period.

### Data sharing

#### National and international collaborators

The study investigators may contribute aggregate and de-identified individual patient data to national and international collaborations whose proposed use of the data has been ethically reviewed and approved by an independent committee and following the signing of an appropriate research collaboration agreement.

#### Data availability statements for research publications

Following the ethical obligations for data sharing outlined by the International Committee for Medical Journal Editors [15], the following data sharing arrangements will be permitted:

*After article publication, the following study data will be made available to researchers from a recognised research institution whose proposed use of the data has been ethically reviewed and approved by an independent committee*.

⍰ *Individual participant data that underlie the results reported in the article, after de-identification (text, tables, figures, and appendices)*
⍰ *The study protocol, statistical analysis plan, analytic code*

*Proposals should be directed to the chief investigator at lisa*.*hui@unimelb*.*edu*.*au. To gain access, data requestors will need to sign a data access agreement with the University of Melbourne*.

## DISCUSSION

This project is the first clinician-led, multi-centre maternity data collection system for metropolitan Melbourne. It complements the state-wide government maternity data, with the major benefits of more timely and flexible reporting of outcomes and granular detail on emerging areas of concern. This resource will support data-informed hospital pandemic or post-pandemic responses through to the end of 2022.

Overall, the quality of the Victorian perinatal data collection is considered high [16]. However, there are limitations to the data collection methods, including missing data on planned home births, private hospital births, and COVID-19 infection rates in pregnancy. There are also hospital-specific variations to the many data collection fields, requiring manual recoding and collation before indicators can be generated.

There are potential risks related to data security. However, as no personally-identifying information will be collected in this study, there is no possibility of breaches of privacy or confidentiality. The risks to the successful conduct of the study include inadequate health service resources to collect and submit the data or insufficient study resources to clean and analyse data on a continuous basis. We have designed this study to leverage existing routine data reporting tools to minimise the burden on health services. We have also obtained philanthropic and University funding to support the central data management and report generation (see Funding declarations).

## SIGNIFICANCE

This collaboration will create a clinician-led resource for data-informed health policy in maternity care. The statistical power of this data collection for rare outcomes such as stillbirth is enhanced by its large sample size and the complete public hospital coverage of the Australian city most affected by pandemic restrictions in 2020. It seamlessly complements the existing department of health state-wide data collection without creating redundancies and onerous workloads for hospital data managers. The system benefits from the advantages of being more agile than existing data sources. The active monthly data collection also allows rapid adjustment of reported indicators to address clinical concerns as they emerge. It is anticipated that this resource will be of enduring value to clinical leaders in maternity health services beyond the peak of the COVID-19 pandemic.

## Data Availability

Data availability statements for research publications: Following the ethical obligations for data sharing outlined by the International Committee for Medical Journal Editors [15], the following data sharing arrangements will be permitted:
After article publication, the following study data will be made available to researchers from a recognised research institution whose proposed use of the data has been ethically reviewed and approved by an independent committee.
⍰ Individual participant data that underlie the results reported in the article, after de-identification (text, tables, figures, and appendices)
⍰ The study protocol, statistical analysis plan, analytic code
Proposals should be directed to the chief investigator at. To gain access, data requestors will need to sign a data access agreement with the University of Melbourne.

## DECLARATIONS

### Ethics approval and consent to participate

Ethics approval for the CoMaND project has been obtained from the Austin Health (HREC/64722/Austin-2020) and Mercy Health (ref. 2020-031). As this is a retrospective review of de-identified health information, there was no requirement for individual consent, or a waiver of consent. This study is registered as an observational study with the Australian New Zealand Clinical Trials Registry (ACTRN12620000878976).

### Consent for publication

All authors have given consent for publication of this manuscript.

## Acknowledgments

We thank Wanyu Chu (Data analyst at the Health Services group, Population Health theme, Murdoch Children’s Research Institute) for supplying the COVID restriction timeline for supplemental file 1. We also acknowledge the contribution of the following hospital staff for their assistance in setting up the data collection within their respective health services: Tania Fletcher (Mercy Health), Michelle Knight (Monash Health), Lynne Rigg (Royal Women’s Hospital), Abby Monaghan (Northern Health), Lee-Anne Lynch (Western Health), Pauline Hamilton (Eastern Health), and Roshanee Perera (Peninsula Health).

## Availability of data and materials

NA

## FINANCIAL DISCLOSURE AND CONFLICTS OF INTEREST

BWM reports consultancy for ObsEva and received research funding from Ferring and Merck. KRP has received consultancy fees from GSK and Janssen Pharmaceuticals. All other investigators do not declare any financial or competing interests for the overall study.

### Authors’ roles in the CoMaND project

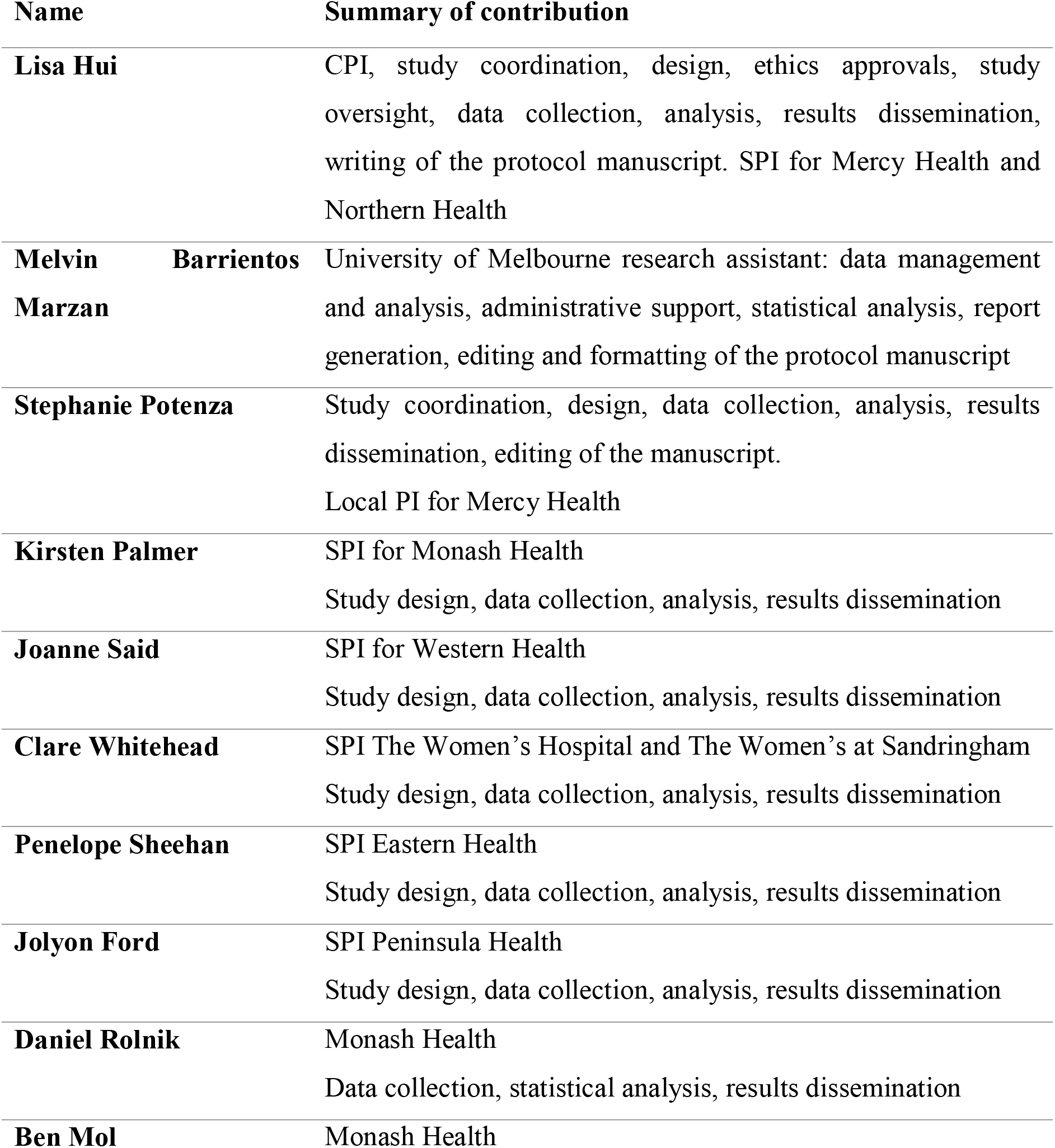
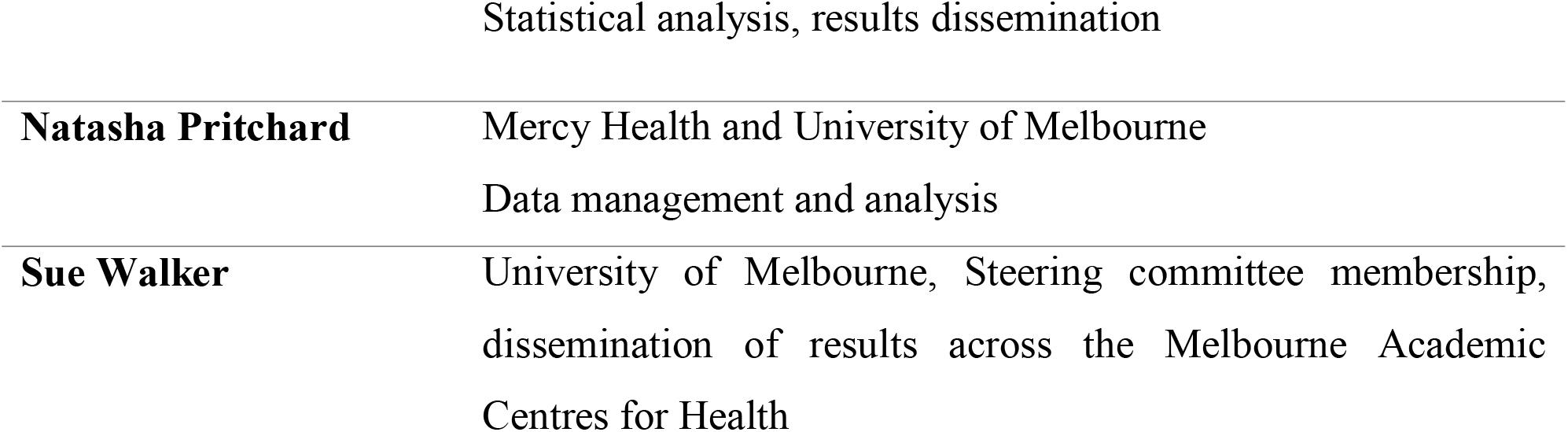

## Abbreviations

ABBREVIATION: TERM
BOS: Birthing Outcome System
BHH: Box Hill Hospital
CCOPMM: Consultative Council of Obstetric and Paediatric Morbidity and Mortality
CoMaND: Collaborative Maternity and Newborn Dashboard
COVID-19: Coronavirus Infectious Disease 2019
CPI: Coordinating Principal Investigator (overall study-level Investigator)
DHHS: Department of Health and Human Services
EDIS: Emergency Department Information System
EH: Eastern Health
FH: Frankston Hospital
HREA: Human Research Ethics Application
HREC: Human Research Ethics Committee
JKWCH: Joan Kirner Women’s and Children’s Hospital
MACH: Melbourne Academic Centre for Health
MCRI: Murdoch Children’s Research Institute
MHW: Mercy Hospital for Women
MMC: Monash Medical Centre
PH: Peninsular Health
PSPI: Perinatal Services Performance Indicators
RWH: Royal Women’s Hospital (Melbourne)
SCV: Safer Care Victoria
SPI: Site Principal Investigator (site-level Investigator)
TAH: The Angliss Hospital
TNH: The Northern Hospital
WMH: Werribee Mercy Hospital
VPDC: Victorian Perinatal Data Collection

