## Supplemental Materials for "A collaborative maternity and newborn dashboard (CoMaND) for the COVID-19 pandemic: a protocol for timely, adaptive monitoring of perinatal outcomes in Melbourne, Australia"

### APPENDIX 1: COVID19 restrictions in Victoria, Australia (timeline)

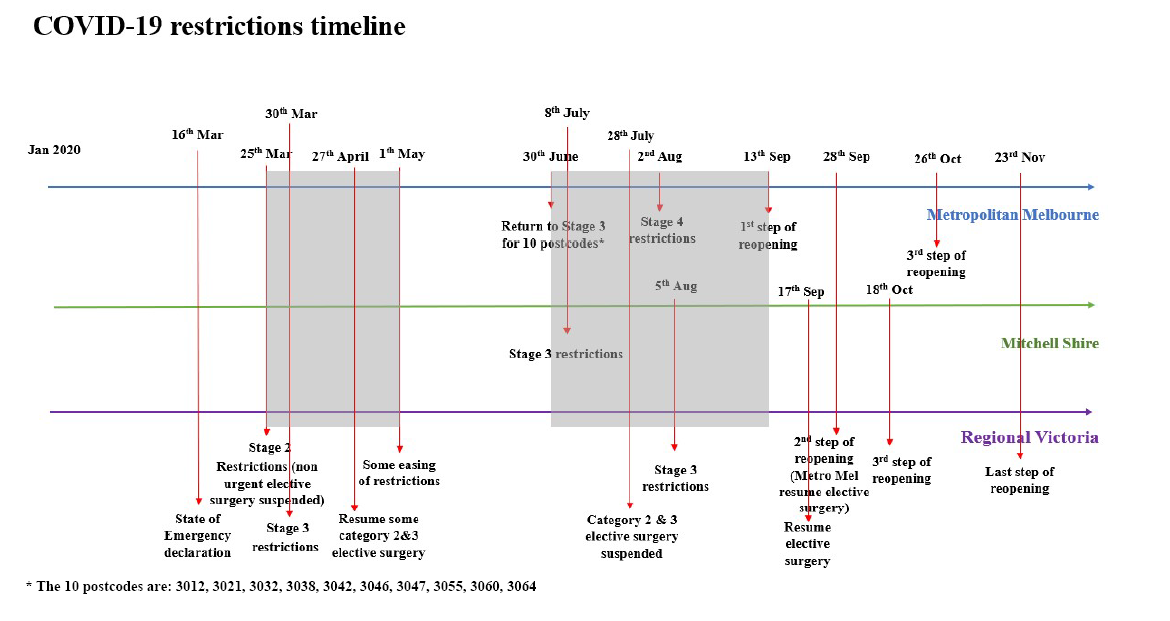

**Summary of Restrictions**

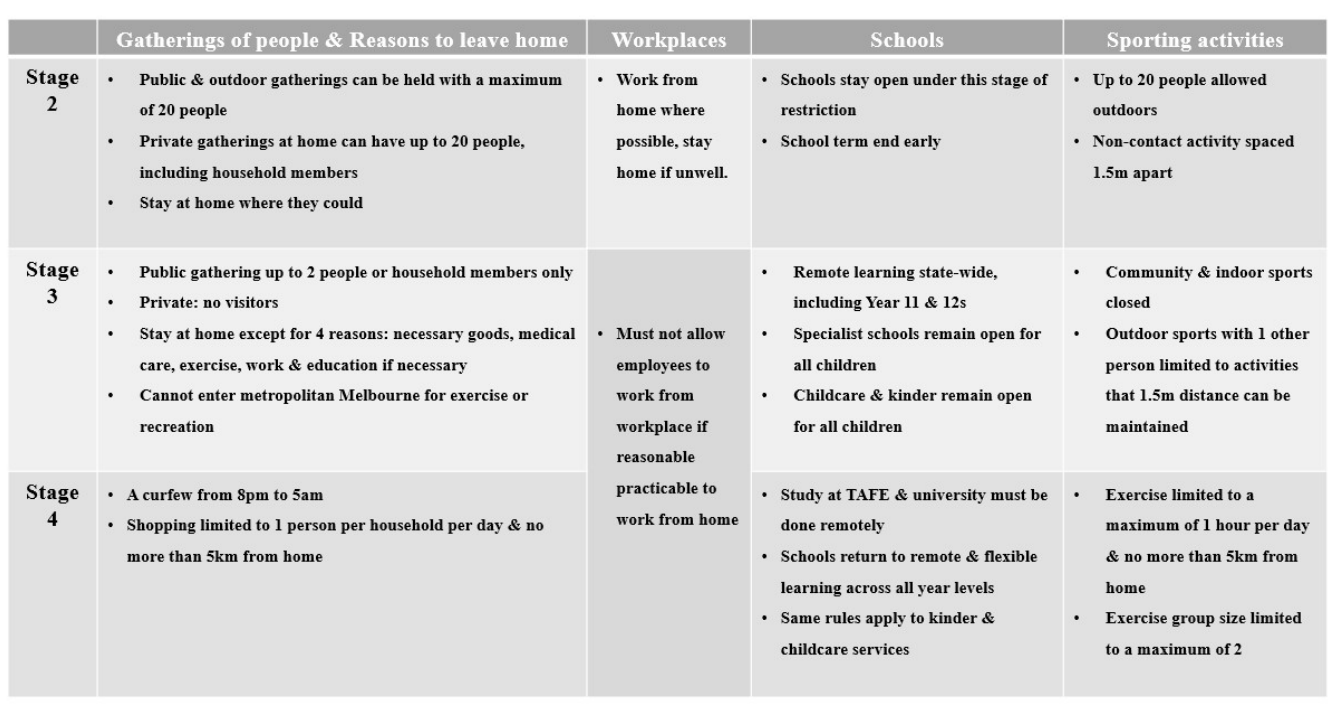

### APPENDIX 2: Tier 2 data collection (customised data fields specific to CoMaND as they appear in the birthing outcomes system)

*NB. Personal identifying fields such as UR, Infant Date of birth, and Mother date of birth will be removed by the local sites prior to submission to study database (grey cells). These fields are required to conduct the data extraction, but are then converted to month and week level data (for infant birth date), and maternal age at delivery (for maternal DOB) before removal. Hospitals that do not use the Birthing Outcomes System (RWH) will extract equivalent data wherever possible.*

1. ID
2. URNo*
3. BabyNum
4. BabyEpisodeID
5. AdmitWeight
6. OnsetLab
7. PlanForVBAC
8. BirthDate*
9. Month
10. Year
11. WeekNum
12. PrimaryBloodLossBirthSuiteEstimated
13. PrimaryBloodLossTheatreEstimated
14. PrimaryBloodLossWardEstimated
15. PrimaryBloodLoss
16. TUCryoprecipitate
17. TUFreshFrozenPlasma
18. TUOther
19. TUPackedRedCells
20. TUPlatelets
21. Transfusion
22. DMGestDiagnosed
23. DMYearDiagnosed
24. AnteFirstVisitGest
25. Gravida
26. AnteCareVisitNumber
27. Parity
28. PlannedBirthType
29. LastSteroid
30. BirthWeight
31. BirthDefectText
32. NeoMorbText
33. Centile
34. BabyScnNicu
35. PlannedTOP
36. ScnNicuLOS
37. Apgar5
38. BirthStatus
39. EstGest
40. BabyGender
41. IntendedPlace
42. IntendedYn
43. IntendedChanged
44. CC01_CriticalCareProvided
45. CC07_CSHysterectomy
46. BMI
47. MmcOtherText
48. Height
49. Weight
50. ObsCompsText
51. Interpreter
52. HdcRequired
53. DOB*
54. MatAge
55. PostCode
56. SmokesAfter20
57. SmokesBefore20
58. BabyLos
59. MotherLos
60. PrevPrimCaesar
61. RefHospital
62. InductMain
63. NoOfSteroidDoses
64. BirthType
65. OpdelMain
66. Presentation
67. DMType
68. Chorionicity
69. Plurality
70. BabyDisHosp
71. BabyDisFeed
72. ExclusiveBF
73. BirthPlace
74. IntendedSpecify
75. IntendedReason
76. Ethnicity
77. PrefLanguage
78. MotherExpectedHomeCare
79. MotherDisDest
80. MotherDisHosp
81. AnteInfluenzaVacc
82. AntePertussisVacc
83. PostPertussisVacc
84. CountryBirth
85. Language
86. AboriginalStatus
87. LatestCareModel
88. InductOther
89. LabAnalgesia
90. BirthAnaes
91. LabComps
92. OpdelOther
93. PeriStatus
94. DMTherapy
95. BirthDefect
96. NeoMorb
97. AttAccoucherRank
98. MmcOther
99. ObsComps
100. PostComps
101. DomViolence
102. PsychosocialFactor
103. Substanc

### APPENDIX 3. Tier 3 data collection: billings for selected MBS item Numbers

| Obstetricians, GPs, Midwives, Nurses Or Aboriginal And Torres Strait Islander Health Practitioners Attendances  These services are for out-of-hospital patients Items introduced 13 March 2020 | | | |
| --- | --- | --- | --- |
| Service | Existing Items *face to face* | COVID-19 Telehealth items *via video-conference* | COVID-19 Telephone items – for when video-conferencing is not available |
| Antenatal Service provided by a Nurse, Midwife or an Aboriginal and Torres Strait Islander health practitioner on behalf of, and under the supervision of, a medical practitioner | 16400 | 91850 | 91855 |
| Postnatal attendance by an obstetrician or GP | 16407 | 91851 | 91856 |
| Postnatal attendance by:  a midwife (on behalf of and under the supervision of the medical practitioner who attended the birth); or  an obstetrician; or  (iii) a general practitioner | 16408 | 91852 | 91857 |
| Antenatal attendance | 16500 | 91853 | 91858 |

| 1. Participating Midwife Attendances. These services are for out-of-hospital patients  Items introduced 13 March 2020 | | | |
| --- | --- | --- | --- |
| Service | Existing Items *face to face* | Telehealth items *via video-conference* | Telephone items *– for when video-conferencing is not available* |
| Short antenatal attendance lasting up to 40 minutes | 82105 | 91211 | 91218 |
| Long antenatal attendance lasting at least 40 minutes | 82110 | 91212 | 91219 |
| Short postnatal attendance lasting up to 40 minutes | 82130 | 91214 | 91221 |
| Long Postnatal attendance lasting at least 40 minutes | 82135 | 91215 | 91222 |

### APPENDIX 4. MBS item numbers – category 5 diagnostic imaging services, group i1 ultrasound, subgroup 5 obstetric and gynaecological

| Item Number | Gestational Age | Plurality |
| --- | --- | --- |
| 55712 | 17-22 weeks | singleton |
| 55721 | > 22 weeks | singleton |
| 55764 | 17-22 weeks | multiple pregnancy |
| 55772 | > 22 weeks | multiple pregnancy |
